# Central cholinergic white matter pathways in prodromal and early manifest Lewy body disease

**DOI:** 10.1101/2025.03.06.25323482

**Authors:** Tamir Eisenstein, Karolien Groenewald, Ludo van Hillegondsberg, Falah Al Hajraf, Tanja Zerenner, Michael A Lawton, Yoav Ben-Shlomo, Ludovica Griffanti, Michele T Hu, Johannes C Klein

## Abstract

**Background and Objectives:** Degeneration of the nucleus basalis of Meynert (NbM) has been reported in Lewy body (LB) disorders. However, while structural changes in the white matter system connecting the NbM to the cerebral cortex have been shown in LB dementia, less is known regarding its vulnerability in prodromal and early manifest patients without cognitive impairment, and its relationship with clinical manifestation and disease progression.

**Methods:** Here, we used diffusion MRI (dMRI) data from the Oxford Parkinson’s Discovery Cohort (OPDC) to examine whether differences in the microstructural integrity of the lateral and medial white matter pathways of the NbM are already evident in prodromal (isolated REM-sleep behaviour disorder (iRBD), n=67), and early manifest (Parkinson’s disease (PD), n=73) LB disease compared to matched controls (n=53). Furthermore, we examined its relationship with baseline and longitudinal cognitive function, and future risk of phenoconverting from iRBD to manifest neurodegenerative disease (PD or dementia with Lewy bodies). Lastly, we examined the potential role of the NbM as a syndrome-specific epicenter in each of the two patient groups by spatially correlating its cortical connectivity profile with cortical atrophy pattern.

**Results:** We found higher microstructural integrity of both pathways to be associated with better verbal fluency performance at baseline (β=3.29-3.52, *p*<0.05). Higher baseline medial pathway integrity was also associated with slower decline in MoCA score over time (β=0.05, *p*<0.05). In addition, higher integrity of both pathways at baseline was associated with reduced future risk of phenoconversion in iRBD (HR<0.51, *p*<0.05). Lastly, we found reduced grey matter volumes in cortical regions that are more anatomically connected to the NbM in iRBD (r=-0.31, *p*<0.05), but not PD (r=-0.08, *p*=0.29), suggesting its potential role in shaping cortical pathology in iRBD. Interestingly, no evidence for differences in NbM pathways integrity between patient cohorts and controls at baseline was observed.

**Conclusions:** Our findings suggest that the NbM white matter system may serve as a non-invasive biomarker, indicating risk for clinical conversion and cortical pathology in iRBD and for baseline and longitudinal cognitive functioning in iRBD and early PD. Hence, it may potentially be used to stratify patients for clinical trials of disease-modifying and neuroprotective therapies.

## Introduction

Cholinergic degeneration and dysfunction have been described in Lewy body (LB) disorders with the most pronounced deficits observed in LB dementia patients^1^. However, while loss of central cholinergic innervation to the cerebral cortex has been proposed as a primary mechanism for cognitive impairment and dementia in LB disorders^2^, previous studies have also shown central cholinergic deficits in LB patients without significant cognitive decline^3–6^. The basal forebrain cholinergic system (BFCS) is the major source of acetylcholine (ACh) to the cortex, hippocampus, and amygdala^7^. BFCS neurons play key roles in shaping the activity of cortical circuits responsible for behavioural and cognitive processing, such as attention, visuospatial skills, and memory^8–11^, which are prominently affected in LB disorders^2,12^. The BFCS is divided into several subregions of which Ch4, which contains the nucleus basalis of Meynert (NbM), provides the main cholinergic innervation to the cerebral cortex. Previous studies have suggested that the NbM may be more susceptible to LB pathology than other regions of the BFCS, as early α-synuclein accumulation and significant atrophy in this region have been observed in LB disease patients^4,13–16^.

Post-mortem studies have shown that NbM’s white matter projections to the cortex travel in two main pathways^7,17^. A lateral pathway travels through the external capsule and uncinate fasciculus, providing cholinergic input to the frontal, parietal, occipital, and temporal cortices, while a medial pathway curves around the rostrum of the corpus callosum, enters the cingulum bundle, and innervates medial cortical regions such as cingulate and retrosplenial cortices. Recently, diffusion-weighted MRI (dMRI) has been utilized to reconstruct these pathways in-vivo, revealing reduced microstructural integrity of these pathways in patients with DLB and Alzheimer’s disease (AD)^18,19^. However, whether the NbM pathways are already affected in early LB disease without dementia, and their potential clinical value is unclear.

LB disorders are characterized by a notably long and diverse prodromal stage which can span decades. While most early prodromal markers of LB disorders are non-specific, a notable exception is isolated REM-sleep behaviour disorder (iRBD)^20^, which is characterized by the loss of muscle atonia during REM sleep phase, leading to dream enactment^21^. Individuals with iRBD are at a high risk for a clinical diagnosis of manifest neurodegenerative disease such as PD or DLB. Previous longitudinal multicenter studies found phenoconversion rates to manifest LB disorder of 6-8% per year, exceeding all known genetic risk, and a long-term risk of phenoconversion in excess of 90%^22–25^, establishing iRBD as one of the strongest clinical markers of prodromal LB disease. However, the time to phenoconversion varies greatly among iRBD patients, ranging from years to even decades after the onset of RBD symptoms. While prominent degeneration of brainstem and peripheral cholinergic nuclei has been reported in iRBD^26^, the extent to which the central cholinergic system in the brain is affected in this prodromal population is less clear.

Therefore, the aims of the present study were four-fold. First, we investigated whether microstructural changes in the NbM white matter pathways are already evident in prodromal and early manifest LB disease (iRBD and early PD, respectively), compared to matched controls. Second, we examined whether the microstructural integrity of the NbM pathways is associated with baseline and longitudinal change in cognitive function. Third, we examined the potential prognostic value of NbM pathways integrity and its association with the risk of future phenoconversion to manifest disease (PD or DLB) in iRBD. Lastly, as being a major source of cortical innervation, we used connectivity-based epicenter analysis to examine whether the NbM could be considered a syndrome-specific epicenter in either iRBD or PD, i.e., a brain region whose connectivity profile may play a central role in brain-wide disease manifestation.

## Materials and methods

### Participants

Data from a total of 73 PD patients, 67 individuals with iRBD, and 53 healthy controls were included in this study from the Oxford Parkinson’s Discovery Cohort (OPDC). The OPDC is a longitudinal observational study aiming to assess progression markers of PD and includes a comprehensive set of clinical and MRI measures (for more details on the cohort see^27^). OPDC participants with valid structural and diffusion MRI data from their baseline visit were included in the study. PD patients were within 3 years of diagnosis. iRBD participants were diagnosed with hospital-based polysomnography and were free of parkinsonism or dementia at recruitment based on objective tests and clinical assessments. All participants underwent the MDS-Unified Parkinson’s Disease Rating Scale (UPDRS) parts III to assess parkinsonian motor features, as well as the Montreal Cognitive Assessment (MoCA) to evaluate general cognitive functioning. Both iRBD and PD patients were free of significant cognitive decline at their baseline assessment. Participants’ demographics are summarized in **Table 1**. The current project was approved by the Research Ethics Board of the University of Oxford, and all participants provided written informed consent according to the Declaration of Helsinki.

**Table 1.**
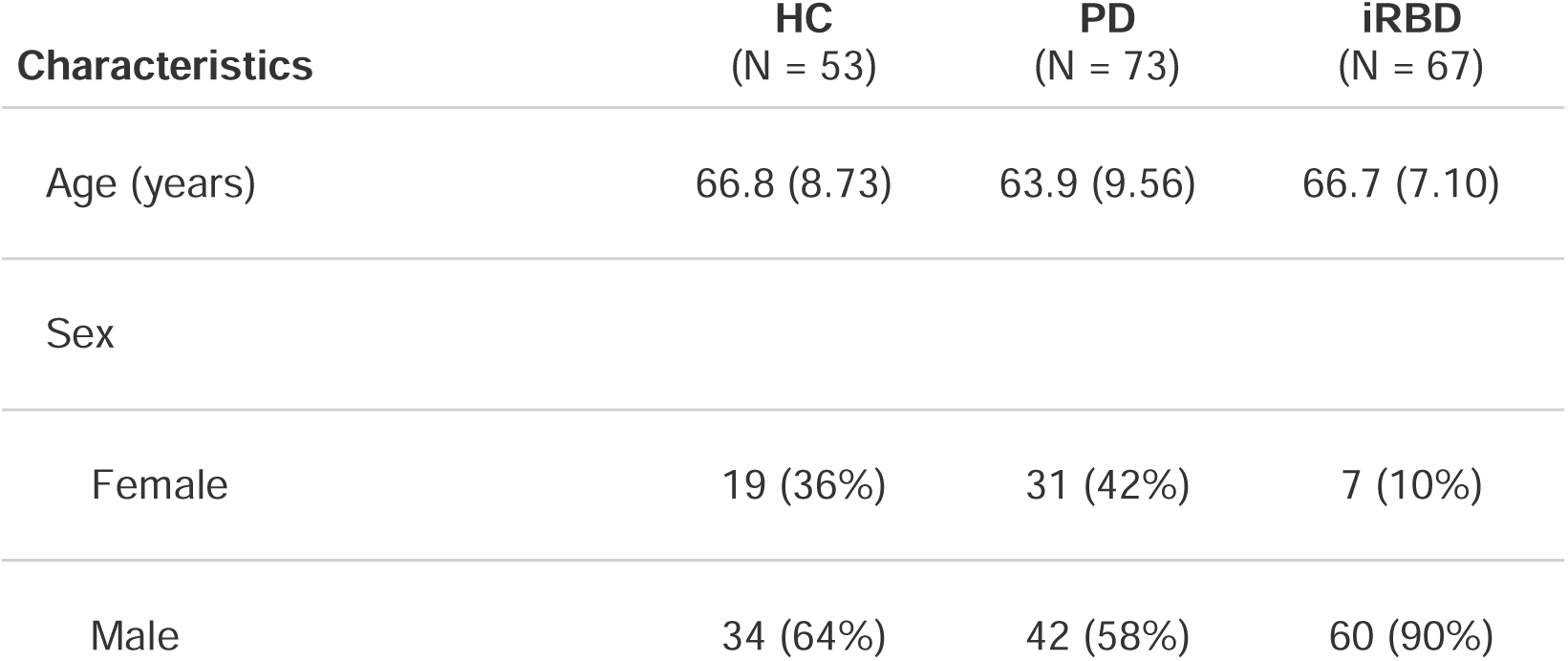
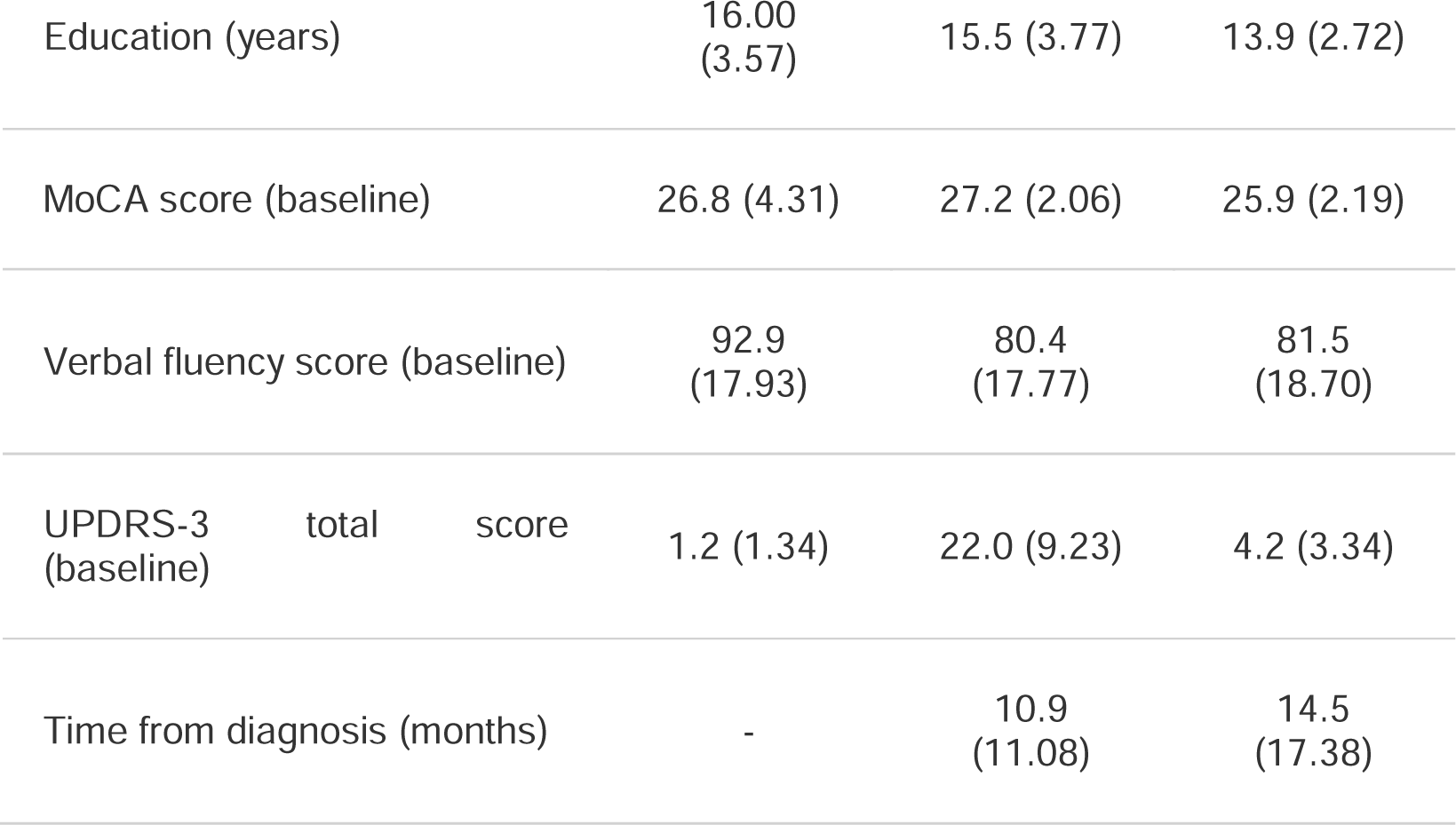
Demographic, cognitive, and clinical characteristics of study groups (mean±SD are presented for continuous variables and proportions for categorical variables)

### MRI acquisition

Data acquisition in the OPDC was performed at the Oxford Centre for Clinical Magnetic Resonance Research (OCMR) using a 3T Trio Siemens MRI scanner equipped with a 12-channel coil. T1-weighted images were obtained using a 3D magnetization prepared-rapid acquisition gradient echo (MPRAGE) sequence (192 axial slices, flip angle 8°, 1 x 1 x 1 mm^3^ voxel size, echo time/repetition time/inversion time = 4.7 ms/2040 ms/900 ms). Total acquisition time 5:56 min. Diffusion MRI data were acquired with an EPI sequence (2 x 2 x 2 mm^3^ voxel size, echo time/repetition time/inversion time = 94 ms/9300 ms, one shells of b-values = 1000 s/mm^2^, 60 unique diffusion gradient directions and 5 b0 images). Total acquisition time 11:11 min.

### Diffusion MRI analysis

The OPDC dMRI data preprocessing was conducted using FSL (FMRIB Software Library https://fsl.fmrib.ox.ac.uk/fsl/fslwiki), and included correction for eddy currents and head motions^28^, skull stripping using the Brain Extraction Tool (BET), and EPI distortion correction using fieldmaps. The percentage of slices identified as outliers across all participants was 0.4±0.39% (mean±SD). Diffusion tensor models were then fitted using DTIfit of the FSL Diffusion Toolbox. Next, tract-based spatial statistics (TBSS) workflow^29^ was utilized to non-linearly register fractional anisotropy (FA) and mean diffusivity (MD) maps into standard MNI space.

### Reconstructing the white matter pathways of the NbM

In order to create templates of the NbM white matter pathways we utilized the preprocessed ultra-high 7T dMRI data of 176 healthy young adults, available in the public release of the Human Connectome Project (HCP) Young Adult study^30,31^ (https://www.humanconnectome.org/study/hcp-young-adult). By using the 7T HCP dMRI data we aimed to base the reconstruction on a high quality-high resolution independent dataset, with better ability to resolve crossing fibres, in order to get a more accurate spatial pattern of the NbM white matter system. Details of the 7T diffusion and T1w image acquisition and preprocessing protocols are provided in the HCP reference manual (https://humanconnectome.org/study/hcp-young-adult/document/1200-subjects-data-release), and the *Supplementary Information*.

Following preprocessing, probabilistic fibre tracking was performed on the preprocessed HCP data with FSL’s ProbtrackX2^31,32^ by generating 5000 random samples from right and left NbM seed region-of-interest (ROI) in MNI space. The NbM seed ROIs were created using the Ch4 masks from the probabilistic cytoarchitectonic map of the Basal Forebrain (v4.2)^33,34^ (**Figure 1a**). A threshold of 50% was applied to probabilistic NbM masks to create the starting region for tractography, following the previously suggested threshold for realistic NbM volume estimation^35^. Probabilistic tractography was guided by several regions of interest based on previous studies^7,18,36^. More details on the process can be found in the *Supplementary Information*. The final resulting unilateral templates of each pathway were then added to create the final bilateral templates of either the medial or lateral pathways (**Figure 1a**).

**Figure 1.**
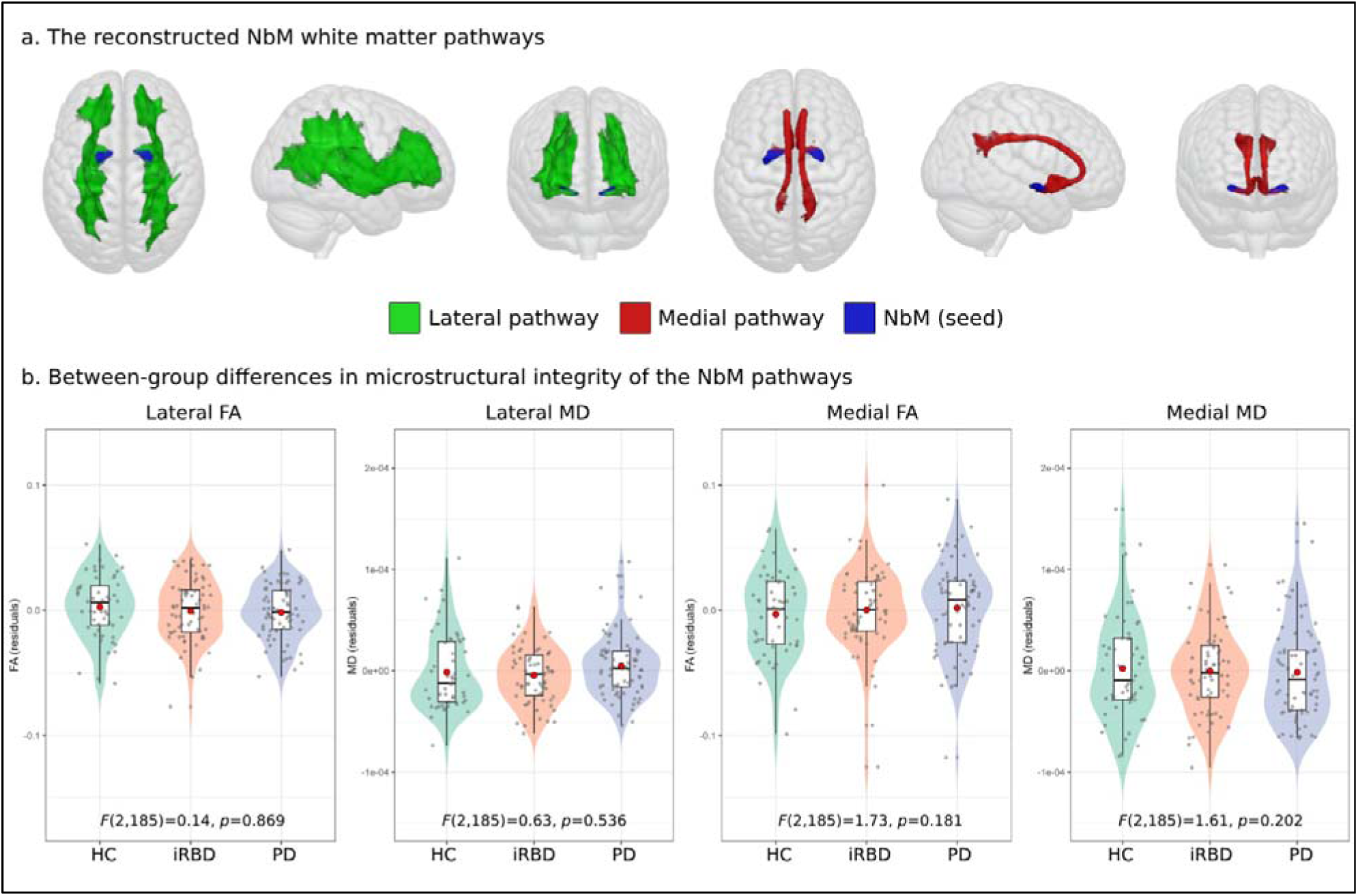
(a) The reconstructed lateral (green) and medial (red) NbM pathways, seeded from the NbM (blue). (b) No between-group differences in FA or MD of either the lateral or medial pathways were found between the study groups. Red circles within boxplots represent group mean.

### Extraction of microstructure indices

To examine the microstructural integrity of the lateral and medial NbM pathways, we transformed the final bilateral pathways templates from MNI to native space for each of the OPDC participants and extracted the averaged FA and MD from the diffusion tensor model’s FA and MD maps of each OPDC participant. Given that the overall (negative) correlation between the FA and MD values across all participants was high (*r*=-0.85), we performed principal component analysis (PCA) on each pathway’s FA and MD metrics to create a single microstructural integrity for that pathway represented by the resulting first PCA component (PC1), which served as the microstructural integrity measure for that pathway in all subject-level analyses. More details are provided in the *Supplementary Information.* Due to potential multicollinearity between the PC1 of the lateral and medial pathways (*r*>0.6), we examined each pathway in a separate model and did not add both measures into the same models.

### NbM grey matter (GM) volume assessment

As previous works demonstrated its vulnerability in Lewy body disorders^14,15^, we also quantified the grey matter volume of the NbM in order to assess the specificity and added value of the white matter component of this system. Measurements of the NbM GM volume were derived from each participant’s T1-weighted MR image using the Computational Anatomy Toolbox (CAT12, https://neuro-jena.github.io/cat/)^37,38^ implemented in Statistical Parametric Mapping (SPM12) software. Default CAT12 preprocessing steps were used to process the raw T1-weighted images and are further detailed in the *Supplementary Information*. The GM volumes of the right and left NbM were extracted from the preprocessed GM images of each participant using the NbM seed ROIs described above. In addition to NbM volume, we also extracted the total intracranial volume (TIV) of each participant to control for differences in head size. The anatomical images were visually inspected and excluded if the image quality rating (IQR) score, derived from CAT12 pipeline, was below 70%. The IQR is a composite measurement integrating several metrics of image quality into a single value ranging from 0 to 100 (i.e., the higher the score, the better the image quality). The final NbM volume metric was computed as the individual residual scores resulting from a linear regression model with NbM volume as the dependent variable and TIV and IQR as explanatory variables.

### Cognitive assessment

In addition to the global cognitive functioning evaluated with the MoCA test, participants also underwent phonemic and semantic verbal fluency tests^39^. Both the MoCA and the verbal fluency tests were evaluated at baseline as well as during eight follow-up visits, with ∼18 months interval between each visit within the iRBD and PD groups. MoCA scores were adjusted for education level as previously suggested^40^. The phonemic and semantic verbal fluency scores were summed within each visit to create a single total verbal fluency score measure. **Supplementary Table 1** summarizes the number of participants with MoCA/verbal fluency data at each visit.

### Phenoconversion from iRBD to defined neurodegeneration

In the current study we have focused on the two major types of phenoconversion among iRBD patients (i.e., to either PD or DLB) and excluded participants who were diagnosed with other neurodegenerative disorder (e.g., multiple systems atrophy). Diagnosis of iRBD to PD or DLB phenoconversion in the OPDC was performed using standard diagnostic criteria^41,42^ applied by trained neurologists assessing each patient longitudinally with a structure series of assessments and examination, with carer report as appropriate.

### Disease-epicenter analysis

To examine the role of the NbM as a potential origin of disease-related structural deficits in iRBD and PD, we conducted an epicenter analysis by spatially correlating the healthy NbM-cortical structural connectivity profile (as derived from the HCP dataset) with the syndrome-specific patterns of cortical atrophy in iRBD and PD from the OPDC cohort^43,44^. Regardless of its atrophy level, a region could be considered a potential epicenter if it is (i) strongly connected to other high-atrophy regions and (ii) weakly connected to low-atrophy regions^43,44^.

#### NbM-cortical structural connectivity profile

To create the healthy NbM-cortical structural connectivity profile we again ran the probabilistic fibre tracking as detailed above using the ProbtracxkX2’s network mode (--network option) to quantify the number of streamlines seeded from either the left or right NbM and 34 cortical regions in each hemisphere using the Desikan-Killiany atlas in MNI space^45^. Streamlines seeded from the NbM to the cortical regions were classified to the lateral or medial NbM pathways as previously suggested and described^7,17,46^ (see *Supplementary Information* for more details).

#### Syndrome-specific cortical atrophy profile

To create the syndrome-specific cortical atrophy profile, a w-score approach was used to account for the expected effects of age, sex, head size and IQR on the structural brain features^47–49^ (see *Supplementary Information* for more details). The w-scores are similar to Z-scores, but are also adjusted for specific covariates (i.e., age, sex, head size and IQR in this case), therefore, representing the normal deviation of the iRBD/PD patient’s GM volume relative to the value expected in the control group (i.e., the OPDC healthy controls) given the patient’s covariates values. From the w-score map of each patient, we extracted the mean w-scores for the 68 cortical regions in the Desikan-Killiany atlas^45^, and then averaged the scores of each region across patients in either the iRBD or PD groups to create the syndrome-specific (i.e., iRBD or PD) cortical atrophy profile (**Figure 5a**). Negative w-score indicated worse/lower score compared to normal expected values accounting for the covariates (i.e. more atrophy/less GM volume).

#### Spatial correlation between connectivity profile and atrophy profiles

Spearman’s rank correlation was used to correlate the spatial patterns of the healthy NbM connectivity profile with each patient group’s cortical atrophy pattern. Since the intrinsic spatial smoothness in two given brain maps may inflate the significance of their spatial correlation, we assessed statistical significance of these correlations using spin permutation tests^43,50^ (see *Supplementary Information* for more details). The original correlation coefficients were then compared against the null distributions determined by the sampling of correlation coefficients with *P*_spin_<0.05 considered statistically significant.

### Statistical analysis

Statistical analyses and visualizations were performed using R version 4.4.0 (https://www.r-project.org/) and the ENIGMA TOOLBOX in Python (https://enigma-toolbox.readthedocs.io/en/latest/index.html). Between-group differences in FA and MD values of the lateral and medial pathways between the three study groups were tested using analysis of covariance (ANCOVA) controlling for age, sex, years of education, and NbM volume. Examination of the relationship between microstructural integrity and cognitive metrics at baseline was conducted using multiple linear regression, controlling for the same covariates. Both main effects and group-based interactions (pathway microstructure x group) were tested. If the interaction term resulted in a *p*-value>0.05 it was removed from the final model for that measure^51^. To examine the relationship between baseline microstructural integrity and longitudinal change in MoCA/verbal fluency performance over time at follow-up visits, we ran a linear mixed model analysis with random intercepts and random slopes using the *‘lme4’* package in R. Cognitive scores at each visit were the dependent variable and participants’ ID the random effect. Time was defined as the time in years from diagnosis of iRBD/PD to each specific visit. Each model included age, sex, years of education, and NbM volume as covariates. To examine whether NbM pathways’ microstructural integrity was related to the extent of change over time, an interaction term of Time x Microstructural integrity was added to each model.

All analyses assessing the relationship between microstructural integrity and cognition were performed on the patient participants only (controls were not included). To test the association between baseline pathways integrity and the risk of phenoconversion in iRBD, we performed Cox proportional hazards regression analyses using the *‘survival’* package in R. Sex was not included in the models due to the very high proportion of males in this group (∼90%). Due to the number of iRBD participants phenoconverting to either PD (n=12) or DLB (n=5) in our cohort, phenoconversion in the models was defined as either DLB or PD diagnosis, i.e., collapsed across both conditions. Time of follow-up in the models was defined as the time interval in months between baseline visit and last visit for patients who did not convert and between baseline visit and date of phenoconversion diagnosis among converted patients. Only patients who had at least one follow-up clinical evaluation after their baseline visit were included in these analyses. Cases were censored when phenoconversion was clinically diagnosed or at their last visit. Schoenfeld residuals method was used to verify that the assumption of proportional hazards was not violated.

## Results

### Demographics

The main demographics and clinical features of the study groups are presented in **Table 1**.

### Between-group comparison of NbM white matter pathways integrity

The lateral and medial NbM tracts reconstructed using the HCP dataset are shown in **Figure 1a**. One-way ANOVA did not reveal any significant differences between the groups in FA or MD of either the lateral or the medial pathways of the NbM after controlling for age, sex, years of education, and NbM volume (lateral-FA: *F*(2,185)=0.14, *p*=0.869; medial-FA: *F*(2,185)=1.73, *p*=0.181; lateral-MD: *F*(2,185)=0.63, *p*=0.536; medial-MD: *F*(2,185)=1.61, *p*=0.202, see **Figure 1b**).

### NbM pathways integrity and cognitive function at baseline

#### MoCA

We first examined whether the two patient groups differed in general cognitive functioning as reflected by the MoCA test. The iRBD group demonstrated significantly lower MoCA score at baseline visit compared to the PD patients (β=-1.07 [95% confidence interval (CI) -1.85, - 0.28], *p*=0.008), when age, sex, and years of education were controlled for. Weak relationships were found between baseline MoCA scores and either the lateral (β=0.13 [95% CI -0.18, 0.44] *p*=0.402) or the medial (β=0.02 [95% CI -0.31, 0.35], *p*=0.910) pathways, controlling for age, sex, years of education, group, and NbM volume (**Figure 2a**). No microstructure integrity x group interactions were observed (lateral: *p*=0.623; medial: *p*=0.650).

**Figure 2.**
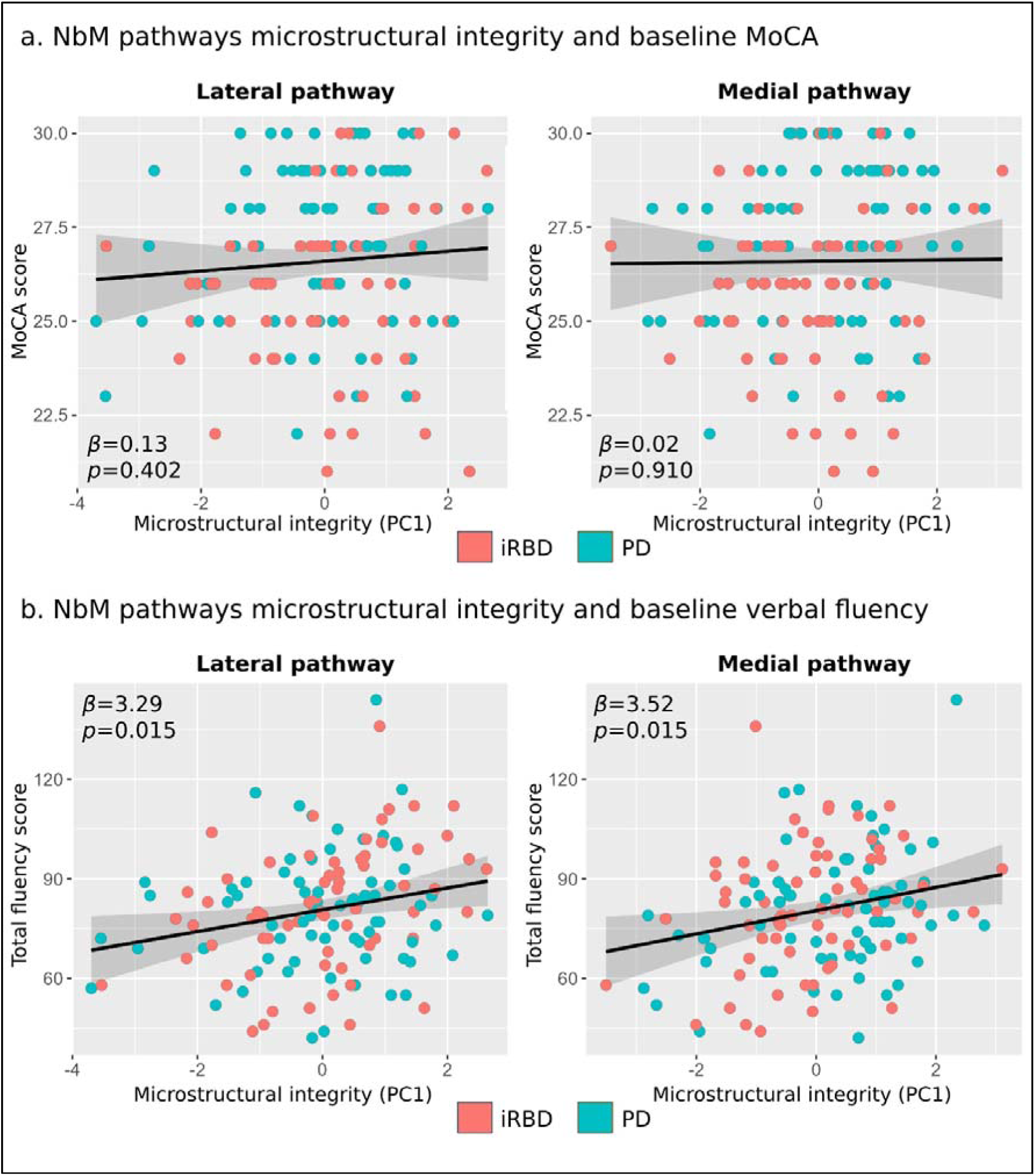
NbM pathways microstructural integrity and cognitive function in prodromal and early Lewy body disease at baseline. (a) No relationship between NbM pathways microstructural integrity and MoCA performance on baseline. (b) Higher lateral and medial pathways integrity at baseline were positively associated with baseline verbal fluency performance.

#### Verbal fluency

In contrast to the MoCA scores, the groups were not statistically different in verbal fluency performance (β=3.05 [95% CI -3.73, 9.83], *p*=0.376), controlling for age, sex, and education. When examining the association between NbM pathway microstructural integrity and verbal fluency performance at baseline visit, we found higher fluency score to be associated with higher microstructural integrity of the lateral pathway (β=3.29 [95% CI 0.66, 5.93], *p*=0.015), and the medial pathway (β=3.52 [95% CI 0.71, 6.33], *p*=0.015), controlling for group, age, sex, education, and NbM volume (**Figure 2b**). No microstructure integrity x group interactions were observed (lateral: *p*=0.094; medial: *p*=0.983).

### NbM pathways integrity and change in cognitive function over time

#### MoCA

MoCA performance was found to significantly decline over time, controlling for age, sex, group, and education (β=-0.11 [95% CI -0.17, -0.05], *p*<0.001), with no time x group interaction (*p*=0.893).

Higher microstructural integrity of the medial pathway was found to be associated with slower decline in MoCA performance over time (**Figure 4a**). We found significant time x microstructural integrity interaction for the medial pathway (β=0.05 [95% CI 0.01, 0.10], *p*=0.026), but not the lateral pathway (β=0.03 [95% CI -0.02, 0.08], *p*=0.299), with no evidence of a 3-way interaction effect when also adding group to the interaction term (medial: *p*=0.931, lateral: *p*=0.933).

**Figure 3.**
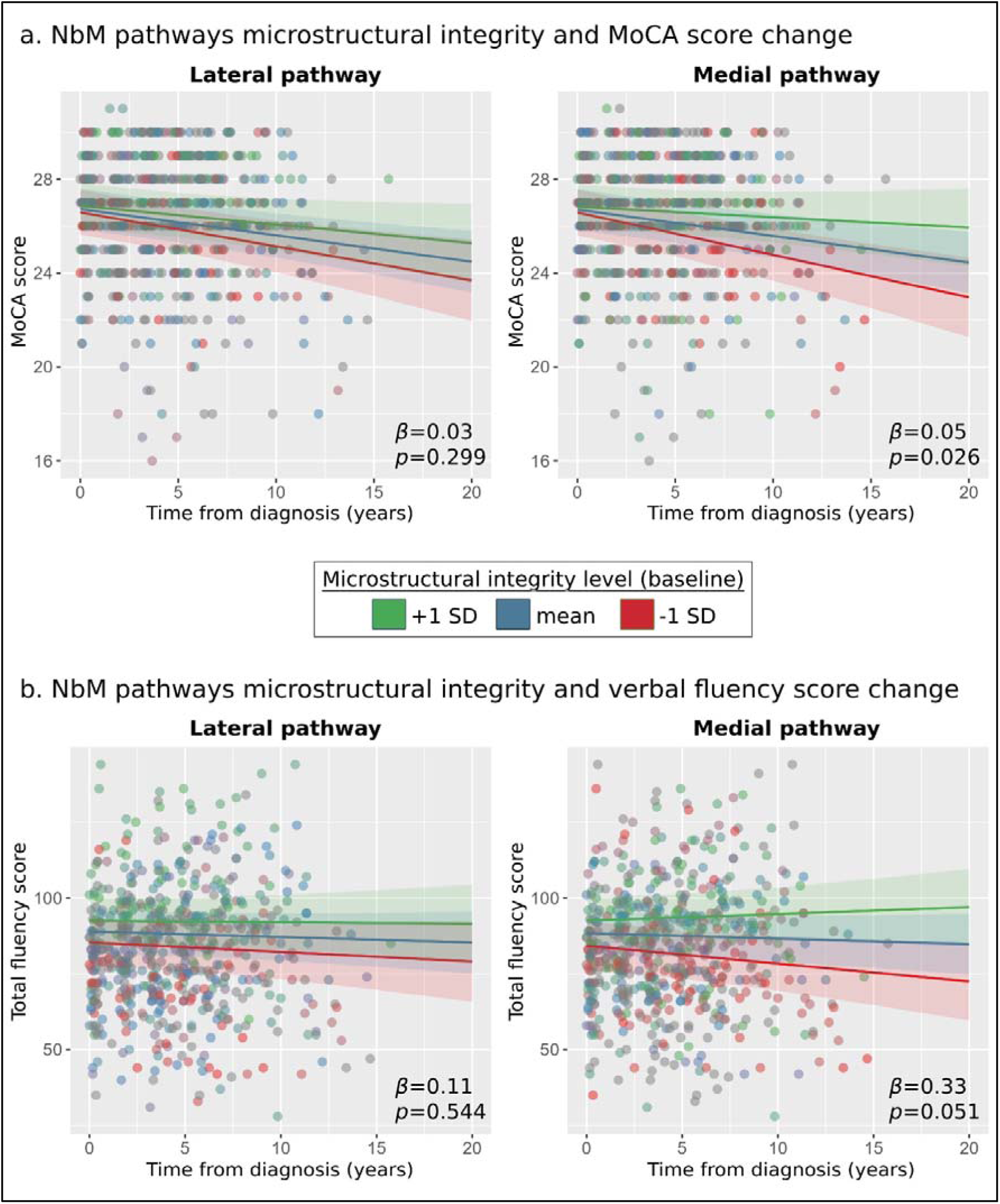
NbM pathways microstructural integrity at baseline and change over time in cognitive function in prodromal and early Lewy body disease. (a) Change in MoCA score over time with a significant time x baseline microstructural integrity interaction for the medial pathway (right panel), suggesting higher medial pathway integrity at baseline is associated with slower decline in MoCA performance over time. (b) Change in verbal fluency score over time, as a function of the lateral and medial pathways integrity at baseline. For visualization purposes of the interaction between cognitive change over time and baseline microstructural integrity, three different slopes representing three levels of baseline microstructural integrity are presented (green = 1 standard deviation (SD) above the mean at baseline, blue=mean at baseline, red = 1 standard deviation below the mean at baseline).

**Figure 4.**
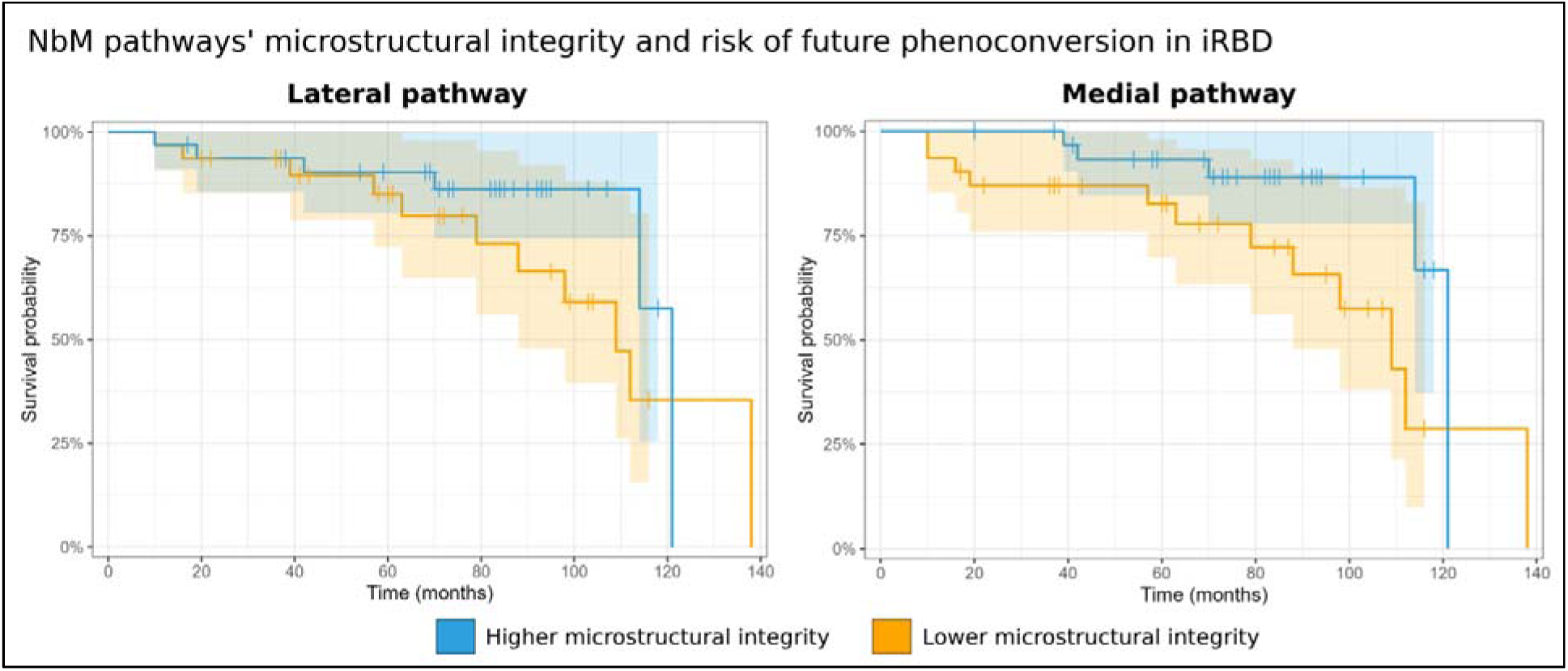
NbM pathways microstructural integrity and risk of future phenoconversion in iRBD. For visualization purposes, we show here the survival probabilities of phenoconverting to PD/DLB in iRBD given higher or lower microstructural integrity at baseline, based on a median split of each pathway’s microstructural values adjusted for age at baseline, time from diagnosis at baseline, education, NbM volume at baseline, and UPDRS-3 and MoCA at baseline (i.e., median splitting the resulting adjusted values/the residuals).

#### Verbal fluency

Interestingly, verbal fluency performance did not significantly change over time, controlling for age, sex, group, and education (β=-0.02 [95% CI -0.05, 0.02], *p*=0.362), with no time x group interaction (*p*=0.110).

While higher microstructural integrity of the medial pathway at baseline visit was found to be associated with the extent of change in MoCA performance over time (**Figure 4b**), a similar association with longitudinal change in verbal fluency performance did not reach statistical significance at the α level of 0.05 (β=0.33 [95% CI -0.003, 0.65], *p*=0.051). The microstructural integrity of the lateral pathway was not associated with verbal fluency performance over time (β=0.11 [95% CI -0.23, 0.44], *p*=0.544). No statistically significant 3- way interaction effects with group as a moderator variable were found (medial: *p*=0.084, lateral: *p*=0.726).

### NbM pathways integrity and risk of phenoconversion in iRBD

During a mean follow-up of 71.0±31.62 months, 17 patients with iRBD phenoconverted to either PD (n=12) or DLB (n=5). We found 1 standard deviation increase in the lateral pathway integrity at baseline to be associated with 44% lower risk of future phenoconversion, when controlling for age and time from diagnosis at baseline visit, years of education, and NbM volume (HR=0.56 [95% CI 0.32, 0.99], *p*=0.045, **Table 2**). Adding MoCA and UPDRS-III scores at baseline as proxies of symptoms severity to further examine the added value of the lateral pathway’s microstructural integrity to the risk of phenoconversion resulted in a ∼49% reduction in risk of phenoconversion for 1 standard deviation increase (HR=0.51 [95% CI 0.27, 0.93], *p*=0.029, **Table 2**, **Figure 4a**).

**Table 2.**
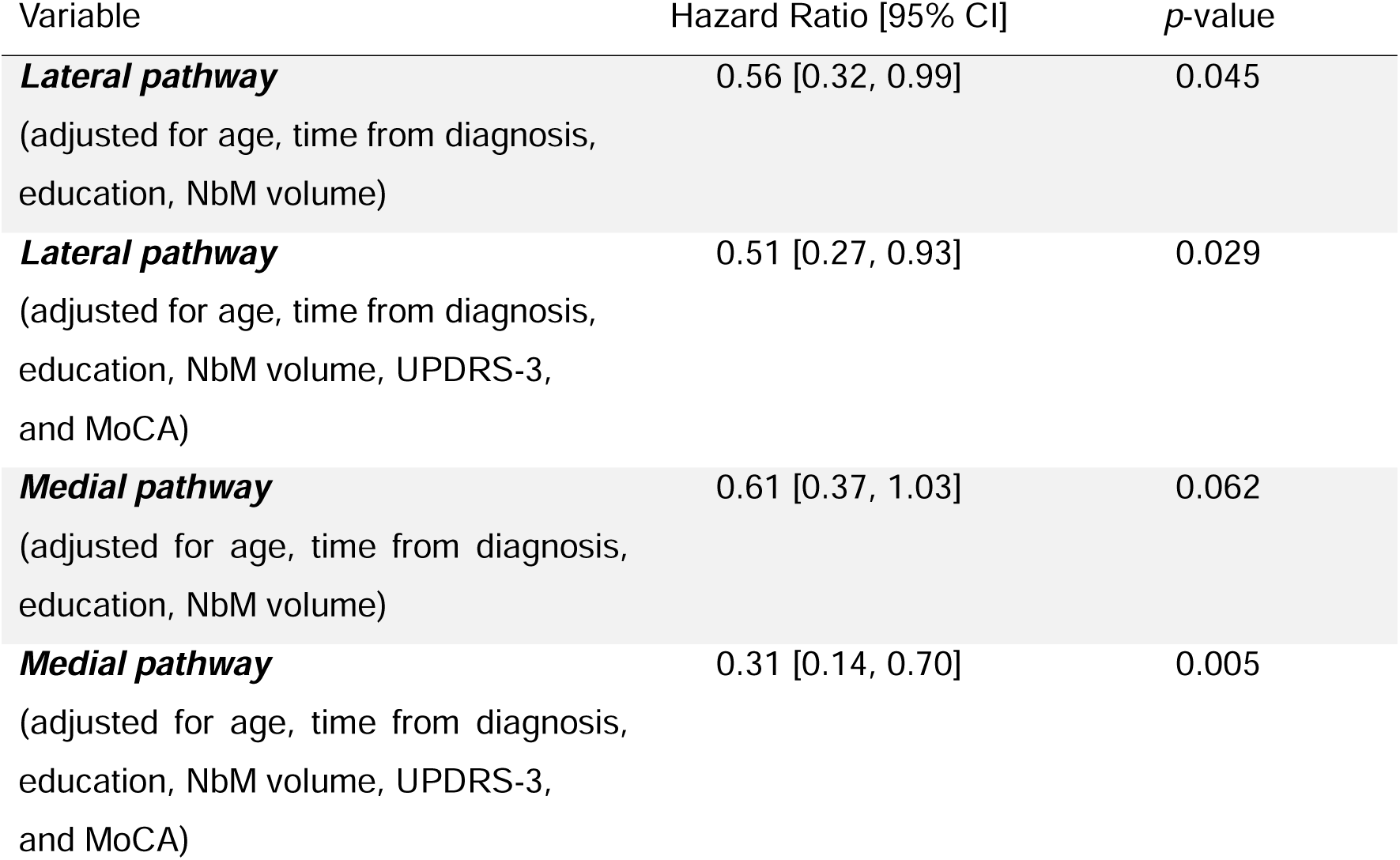
Survival analysis of NbM pathways microstructural integrity and risk of phenoconversion in iRBD patients. Hazard Ratio coefficients are per 1 standard deviation of NbM volume.

| Variable | Hazard Ratio [95% CI] | *p*-value |
| --- | --- | --- |
| **Lateral pathway**<br>(adjusted for age, time from diagnosis,<br>education, NbM volume) | 0.56 [0.32, 0.99] | 0.045 |
| **Lateral pathway**<br>(adjusted for age, time from diagnosis,<br>education, NbM volume, UPDRS-3,<br>and MoCA) | 0.51 [0.27, 0.93] | 0.029 |
| **Medial pathway**<br>(adjusted for age, time from diagnosis,<br>education, NbM volume) | 0.61 [0.37, 1.03] | 0.062 |
| **Medial pathway**<br>(adjusted for age, time from diagnosis,<br>education, NbM volume, UPDRS-3,<br>and MoCA) | 0.31 [0.14, 0.70] | 0.005 |

We also found 1 standard deviation increase in the medial pathway integrity at baseline to be associated with ∼39% lower risk of future phenoconversion, when controlling for age at baseline, time from diagnosis at baseline, years of education, and NbM volume, although consistent with change at α level of 0.05 (HR=0.61 [95% CI 0.37, 1.03], *p*=0.062, **Table 2**, **Figure 4b**). Adding MoCA and UPDRS-III scores at baseline to the model revealed a ∼69% risk reduction for medial pathway microstructural integrity 1 standard increase (HR=0.31 [95% CI 0.14, 0.70], *p*=0.005, **Table 2**, **Figure 4b**).

### NbM as a disease epicenter in iRBD and PD

Lastly, we aimed to examine the potential role of the NbM as a disease epicenter in iRBD and PD by examining the spatial relationship between the structural connectivity profile of the NbM and the cortical atrophy patterns of both patient groups. Both iRBD and PD patients demonstrated a general anterior-to-posterior pattern of cortical volume differences compared to controls, with negative w-scores (i.e., lower volume/higher atrophy) found predominantly in posterior cortices (**Figure 5a**). We found a significant negative spatial correlation between the cortical atrophy profile and the healthy HCP-derived structural connectivity profile of the NbM in the iRBD group (*r*=-0.31, *p_spin_*=0.023), suggesting that cortical regions that are more anatomically connected to the NbM also demonstrated a higher degree of atrophy in this disease group (**Figure 5b**). We did not observe a spatial relationship between atrophy patterns and NbM connectivity patterns in the PD group (*r*=-0.08, *p*=0.293) (**Figure 5b**).

**Figure 5.**
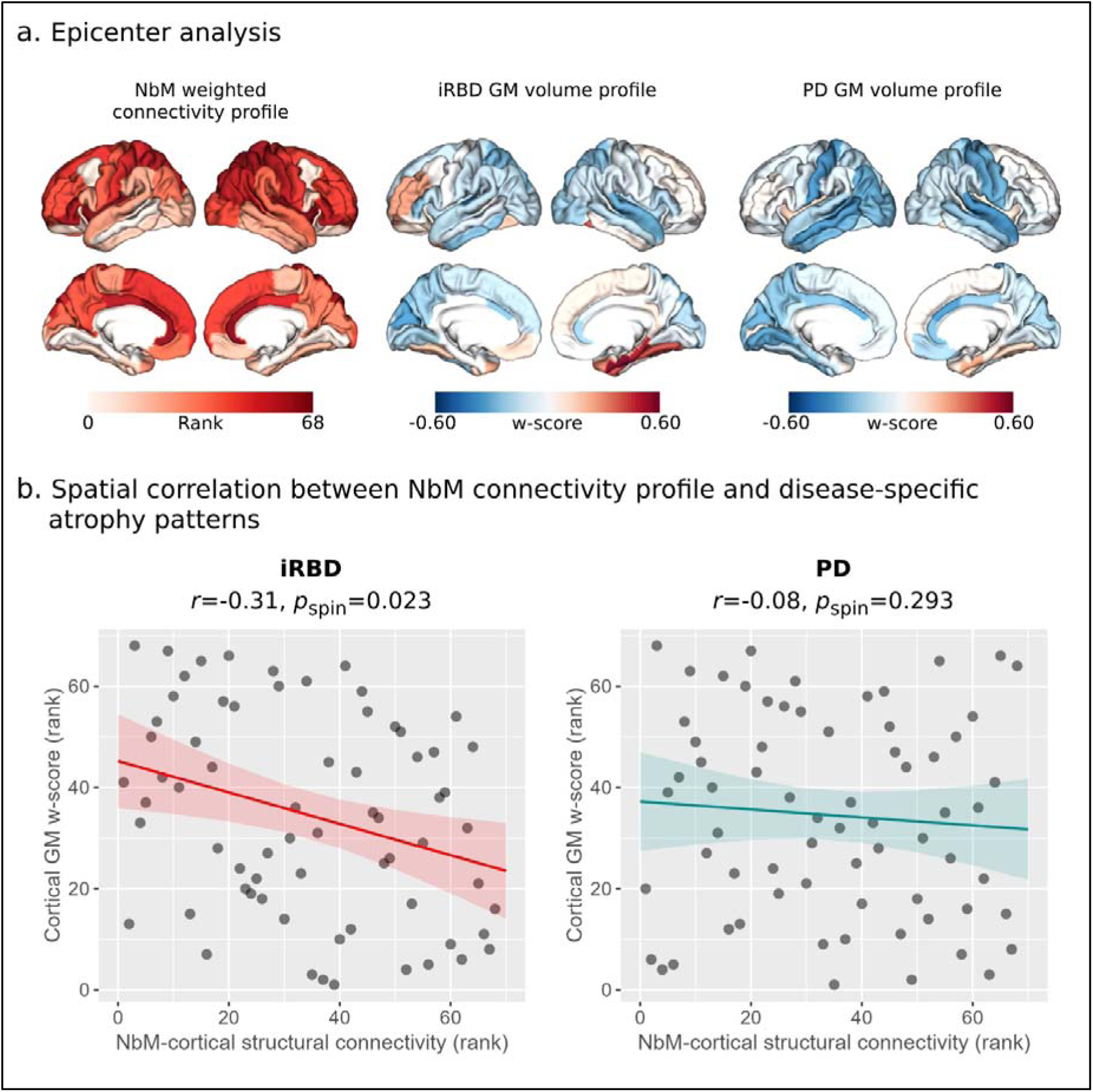
NbM as a potential syndrome-specific epicenter in prodromal and manifest Lewy body disorders. (A) Disease epicenter analysis scheme, based on normative NbM-cortical structural connectivity profile derived from the HCP dataset (left panel, regions with greater weighted connectivity are presented in darker red), and syndrome-specific cortical GM volume patterns in iRBD (middle panel) and PD (right panel), expressed as w-scores relative to the age-matched controls in the OPDC cohort. (B) NbM connectivity profile correlated with atrophy patterns in iRBD (left panel), but not PD (right panel), suggesting its potential role as origin of cortical structural deficits in this prodromal syndrome.

## Discussion

Degeneration of the NbM has been observed in PD patients with and without dementia^4,52^, and is suggested to occur already in patients with iRBD^53^. Despite the early suggested involvement of NbM pathology in LB disorders^16^, there is limited evidence on the white matter system originating from the NbM in prodromal and manifest LB disorders without cognitive decline. The present study aimed to investigate the microstructural integrity of NbM white matter pathways in individuals with iRBD and patient with early manifest PD without cognitive decline, to examine its association with cognitive performance and disease progression (i.e., longitudinal change in cognitive function and phenoconversion), and whether the NbM, through its structural connections with the cerebral cortex, could be considered a disease epicenter in those patient groups, potentially shaping cortical structural deficits.

The NbM cholinergic system has been implicated in cognitive dysfunction in Lewy body disorders, especially in attention, visuospatial, and memory processing^2,9,12^. This study found that the microstructural integrity of NbM pathways is associated with baseline and longitudinal cognitive function in LB disorders patients without cognitive decline. Higher pathways integrity was associated with better baseline verbal fluency. Additionally, increased integrity of the medial pathway at baseline was also associated with milder MoCA decline over time and showed a similar trend for verbal fluency over time.

Previous studies have shown associations between the NbM pathways integrity and better performance across various cognitive tests and populations. Schumacher and colleagues found a link between lateral NbM tract integrity and the Mini Mental State Examination (MMSE) test for global cognition and choice reaction time test across patients with MCI and manifest AD and DLB^18^. Nemy and colleagues found association between NbM tracts integrity and tests of attention and memory in MCI and AD patients, and attention in cognitively normal individuals^19^. However, no association was found with the Cambridge Cognitive Examination-Revised test battery (CAMCOG) performance in PD^54^. The tests in which performance was linked with NbM pathways integrity involve multiple cognitive processes, including attention and memory, which are particularly linked with the cholinergic system. Verbal fluency performance, associated with NbM pathway integrity in this study, also involves multiple cognitive processes such as attention, working and semantic memory, executive functioning and language^39,55^, aligning with the cholinergic system’s role in vast cortical circuits modulation. The lack of association between NbM pathways integrity and baseline MoCA scores observed in the current study, could be due to the relatively limited range of MoCA scores in those patients, who are without significant cognitive deficits. Associations between the general MoCA test and cholinergic markers may become more apparent as disease progresses, especially among early manifest patients without cognitive deficits.

One of our most meaningful findings was the potential predictive value of NbM pathways integrity for disease progression in iRBD as lower integrity was strongly associated with increased risk of phenoconversion. As this association persisted even after rigorous control for disease severity symptoms such as UPDRS-3 and general cognitive functioning, it highlights these pathways potential as independent biomarkers for risk stratification and participants selection in clinical trials of disease-modifying and neuroprotective therapies. While our sample size precluded differentiation between PD/DLB phenoconversion, this limitation may be less consequential than initially apparent. Even large multi-center collaborations have struggled establishing clear distinctions between these conversion types^23^, which likely reflects the fundamental shared similarities in clinical presentation, and the fact that iRBD patients convert to either condition in roughly similar proportions^23^. Our findings align with and extend previous research on the NbM tracts’ predictive value in neurodegenerative progression, as reduced NbM pathways integrity was also found to predict higher risk of dementia onset in MCI patients^18^. Together, these results strengthen the emerging role of NbM pathways integrity as a valuable prognostic marker across different stages of neurodegeneration. In addition, to further support the predictive role that the NbM white matter system may play in iRBD, we also found this region, through its cortical white matter connections, to be associated with cortical atrophy in iRBD, but not PD. It is possible that the cholinergic system plays a more central role in iRBD compared to early manifest PD without cognitive decline, given the high proportion of dementia conversion in this prodromal subtype of LB disorders^23^ and the well-documented vulnerability of this system in Lewy body dementia^9^.

Interestingly, despite the successful NbM pathways reconstruction as previously described^7,17,18,36^, the vulnerability of NbM neurons in LB disorders and the subject-level associations we observed, no significant differences in pathways integrity were found between patients and controls. This partially contrasts with a previous study that found differences in the lateral pathway’s integrity in prodromal and manifest DLB patients^18^. However, those prodromal DLB patients were already presenting with objective cognitive deficits, aligning with the more severe cholinergic deficits reported in Lewy body dementia patients^3,52^. Furthermore, Hepp and colleagues^54^ have also not found differences in NbM tracts intergity between PD patients, either with or without hallucinations, and controls. It is possible that standard DTI metrics such as FA and MD, are not sensitive enough to detect cholinergic white matter loss in prodromal and early manifest patients without cognitive decline, where the expected deficits could be more subtle. More sensitive dMRI techniques, such as multi-shell acquisitions, may be needed to better model microstructural sub-components, address potential partial volume effects, and provide deeper neuropathological insights^56,57^.

### Limitations

Despite iRBD and PD representing distinct phases of Lewy body disease continuum, no significant group-based interactions were found between NbM tracts integrity and clinical or cognitive measures. However, the absence of significant interactions should be interpreted with caution. Detection of interaction effects requires substantially larger sample sizes compared to main effects^58^, suggesting our study may have been underpowered to identify subtle group-based interactions. Additionally, our relatively modest number of iRBD-to-manifest disease converters underscores the need for validation studies with larger converter cohorts to strengthen and extend our findings.

Furthermore, while we used state-of-the-art ultra-high resolution 7T dMRI data to reconstruct the NBM pathways, current in-vivo dMRI methods cannot completely isolate cholinergic fibers from other fiber systems, limiting the ability to draw definitive conclusions about the pure contribution of the actual cholinergic fibres to these pathways.

### Conclusions

Our study reveals that the degree of NbM pathways microstructural integrity holds important clinical significance. NbM pathways integrity correlates meaningfully with both baseline and longitudinal cognitive performance in prodromal early manifest LB patients. Further, our findings may help establish NbM pathways integrity as a promising non-invasive imaging biomarker for predicting disease progression risk to manifest disease in iRBD patients and suggest a potential role of the NbM in shaping disease-related structural cortical deficits. These results point towards a valuable tool for patient stratification in clinical trials focused on disease-modifying and neuroprotective therapies. To strengthen these findings and establish their clinical utility, future research should focus on validating these results in larger cohorts, particularly with expanded samples of phenoconverters.

## Data Availability

OPDC data are available upon reasonable request. Qualified investigators seeking access to de-identified participant data relating to the OPDC may submit their request by means of a formal application to the Oxford Parkinson’s Research Centre (OPDC) Data Access Committee. The application form, protocol, and terms and conditions may be found at opdc.medsci.ox.ac.uk/external-collaborations.

## Acknowledgments

For the purpose of open access, the authors have applied a CC BY public copyright licence to any Author Accepted Manuscript version arising from this submission.

## References

1. Pasquini J, Brooks DJ, Pavese N. The Cholinergic Brain in Parkinson’s Disease. Mov Disord Clin Pract. United States; 2021;8:1012–1026.

2. Aarsland D, Batzu L, Halliday GM, et al. Parkinson disease-associated cognitive impairment. Nat Rev Dis Prim [online serial]. 2021;7:47. Accessed at: 10.1038/s41572-021-00280-3.

3. Klein JC, Eggers C, Kalbe E, et al. Neurotransmitter changes in dementia with Lewy bodies and Parkinson disease dementia in vivo. Neurology. United States; 2010;74:885–892.

4. Liu AKL, Chang RC-C, Pearce RKB, Gentleman SM. Nucleus basalis of Meynert revisited: anatomy, history and differential involvement in Alzheimer’s and Parkinson’s disease. Acta Neuropathol. Germany; 2015;129:527–540.

5. van der Zee S, Kanel P, Gerritsen MJJ, et al. Altered Cholinergic Innervation in De Novo Parkinson’s Disease with and Without Cognitive Impairment. Mov Disord. United States; 2022;37:713–723.

6. Bohnen NI, Kaufer DI, Hendrickson R, et al. Cognitive correlates of cortical cholinergic denervation in Parkinson’s disease and parkinsonian dementia. J Neurol. Germany; 2006;253:242–247.

7. Mesulam MM. Cholinergic circuitry of the human nucleus basalis and its fate in Alzheimer’s disease. J Comp Neurol. United States; 2013;521:4124–4144.

8. Mesulam M. Cholinergic aspects of aging and Alzheimer’s disease. Biol Psychiatry. United States; 2012;71:760–761.

9. Bohnen NI, Grothe MJ, Ray NJ, Müller MLTM, Teipel SJ. Recent advances in cholinergic imaging and cognitive decline-Revisiting the cholinergic hypothesis of dementia. Curr Geriatr reports. United States; 2018;7:1–11.

10. Avery MC, Krichmar JL. Neuromodulatory Systems and Their Interactions: A Review of Models, Theories, and Experiments. Front Neural Circuits. Switzerland; 2017;11:108.

11. Ananth MR, Rajebhosale P, Kim R, Talmage DA, Role LW. Basal forebrain cholinergic signalling: development, connectivity and roles in cognition. Nat Rev Neurosci [online serial]. 2023;24:233–251. Accessed at: 10.1038/s41583-023-00677-x.

12. Kehagia AA, Barker RA, Robbins TW. Cognitive impairment in Parkinson’s disease: the dual syndrome hypothesis. Neurodegener Dis. Switzerland; 2013;11:79–92.

13. Giguère N, Burke Nanni S, Trudeau L-E. On Cell Loss and Selective Vulnerability of Neuronal Populations in Parkinson’s Disease. Front Neurol [online serial]. 2018;9. Accessed at: https://www.frontiersin.org/journals/neurology/articles/10.3389/fneur.2018.00455.

14. Ray NJ, Bradburn S, Murgatroyd C, et al. In vivo cholinergic basal forebrain atrophy predicts cognitive decline in de novo Parkinson’s disease. Brain. England; 2018;141:165–176.

15. Schumacher J, Kanel P, Dyrba M, et al. Structural and molecular cholinergic imaging markers of cognitive decline in Parkinson’s disease. Brain [online serial]. 2023;146:4964–4973. Accessed at: 10.1093/brain/awad226.

16. Braak H, Del Tredici K, Rüb U, de Vos RAI, Jansen Steur ENH, Braak E. Staging of brain pathology related to sporadic Parkinson’s disease. Neurobiol Aging. United States; 2003;24:197–211.

17. Selden NR, Gitelman DR, Salamon-Murayama N, Parrish TB, Mesulam MM. Trajectories of cholinergic pathways within the cerebral hemispheres of the human brain. Brain. England; 1998;121 (Pt 1:2249–2257.

18. Schumacher J, Ray NJ, Hamilton CA, et al. Cholinergic white matter pathways in dementia with Lewy bodies and Alzheimer’s disease. Brain [online serial]. 2022;145:1773–1784. Accessed at: 10.1093/brain/awab372.

19. Nemy M, Dyrba M, Brosseron F, et al. Cholinergic white matter pathways along the Alzheimer’s disease continuum. Brain [online serial]. 2023;146:2075–2088. Accessed at: 10.1093/brain/awac385.

20. Berg D, Borghammer P, Fereshtehnejad S-M, et al. Prodromal Parkinson disease subtypes — key to understanding heterogeneity. Nat Rev Neurol [online serial]. 2021;17:349–361. Accessed at: 10.1038/s41582-021-00486-9.

21. Högl B, Stefani A, Videnovic A. Idiopathic REM sleep behaviour disorder and neurodegeneration — an update. Nat Rev Neurol [online serial]. 2018;14:40–55. Accessed at: 10.1038/nrneurol.2017.157.

22. Iranzo A, Fernández-Arcos A, Tolosa E, et al. Neurodegenerative Disorder Risk in Idiopathic REM Sleep Behavior Disorder: Study in 174 Patients. PLoS One [online serial]. Public Library of Science; 2014;9:e89741. Accessed at: 10.1371/journal.pone.0089741.

23. Postuma RB, Iranzo A, Hu M, et al. Risk and predictors of dementia and parkinsonism in idiopathic REM sleep behaviour disorder: a multicentre study. Brain. England; 2019;142:744–759.

24. Postuma RB, Iranzo A, Hogl B, et al. Risk factors for neurodegeneration in idiopathic rapid eye movement sleep behavior disorder: a multicenter study. Ann Neurol. United States; 2015;77:830–839.

25. Iranzo A, Tolosa E, Gelpi E, et al. Neurodegenerative disease status and post-mortem pathology in idiopathic rapid-eye-movement sleep behaviour disorder: an observational cohort study. Lancet Neurol. England; 2013;12:443–453.

26. Dauvilliers Y, Schenck CH, Postuma RB, et al. REM sleep behaviour disorder. Nat Rev Dis Prim [online serial]. 2018;4:19. Accessed at: 10.1038/s41572-018-0016-5.

27. Griffanti L, Klein JC, Szewczyk-Krolikowski K, et al. Cohort profile: the Oxford Parkinson’s Disease Centre Discovery Cohort MRI substudy (OPDC-MRI). BMJ Open. England; 2020;10:e034110.

28. Andersson JLR, Sotiropoulos SN. An integrated approach to correction for off-resonance effects and subject movement in diffusion MR imaging. Neuroimage. 2015/10/21. 2016;125:1063–1078.

29. Smith SM, Jenkinson M, Johansen-Berg H, et al. Tract-based spatial statistics: voxelwise analysis of multi-subject diffusion data. Neuroimage. 2006/04/21. 2006;31:1487–1505.

30. Van Essen DC, Smith SM, Barch DM, Behrens TEJ, Yacoub E, Ugurbil K. The WU-Minn Human Connectome Project: an overview. Neuroimage. United States; 2013;80:62–79.

31. Warrington S, Bryant KL, Khrapitchev AA, et al. XTRACT - Standardised protocols for automated tractography in the human and macaque brain. Neuroimage [online serial]. 2020;217:116923. Accessed at: https://www.sciencedirect.com/science/article/pii/S1053811920304092.

32. Behrens TEJ, Berg HJ, Jbabdi S, Rushworth MFS, Woolrich MW. Probabilistic diffusion tractography with multiple fibre orientations: What can we gain? Neuroimage [online serial]. 2007;34:144–155. Accessed at: https://www.sciencedirect.com/science/article/pii/S1053811906009360.

33. Zaborszky L, Hoemke L, Mohlberg H, Schleicher A, Amunts K, Zilles K. Stereotaxic probabilistic maps of the magnocellular cell groups in human basal forebrain. Neuroimage. United States; 2008;42:1127–1141.

34. Zaborszky, L., Hoemke, L., Mohlberg, H., Schleicher, A., Amunts, K., & Zilles K. Probabilistic cytoarchitectonic map of Ch 4 (Basal Forebrain) [data set]. Hum Brain Proj Neuroinformatics Platf. Epub 2019.

35. Wang Y, Zhan M, Roebroeck A, De Weerd P, Kashyap S, Roberts MJ. Inconsistencies in atlas-based volumetric measures of the human nucleus basalis of Meynert: A need for high-resolution alternatives. Neuroimage. United States; 2022;259:119421.

36. Nemy M, Cedres N, Grothe MJ, et al. Cholinergic white matter pathways make a stronger contribution to attention and memory in normal aging than cerebrovascular health and nucleus basalis of Meynert. Neuroimage. United States; 2020;211:116607.

37. Gaser C, Dahnke R, Thompson PM, Kurth F, Luders E. CAT – A Computational Anatomy Toolbox for the Analysis of Structural MRI Data. bioRxiv [online serial]. Epub 2023 Jan 1.:2022.06.11.495736. Accessed at: http://biorxiv.org/content/early/2023/11/28/2022.06.11.495736.abstract.

38. Tremblay C, Rahayel S, Vo A, et al. Brain atrophy progression in Parkinson’s disease is shaped by connectivity and local vulnerability. Brain Commun. England; 2021;3:fcab269.

39. Henderson SK, Peterson KA, Patterson K, Lambon Ralph MA, Rowe JB. Verbal fluency tests assess global cognitive status but have limited diagnostic differentiation: evidence from a large-scale examination of six neurodegenerative diseases. Brain Commun. England; 2023;5:fcad042.

40. Nasreddine ZS, Phillips NA, Bédirian V, et al. The Montreal Cognitive Assessment, MoCA: A brief screening tool for mild cognitive impairment. J Am Geriatr Soc. 2005;53:695–699.

41. Szewczyk-Krolikowski K, Tomlinson P, Nithi K, et al. The influence of age and gender on motor and non-motor features of early Parkinson’s disease: initial findings from the Oxford Parkinson Disease Center (OPDC) discovery cohort. Parkinsonism Relat Disord. England; 2014;20:99–105.

42. McKeith IG, Ferman TJ, Thomas AJ, et al. Research criteria for the diagnosis of prodromal dementia with Lewy bodies. Neurology. United States; 2020;94:743–755.

43. Larivière S, Rodríguez-Cruces R, Royer J, et al. Network-based atrophy modeling in the common epilepsies: A worldwide ENIGMA study. Sci Adv. United States; 2020;6.

44. Jiang Y, Palaniyappan L, Luo C, et al. Neuroimaging epicenters as potential sites of onset of the neuroanatomical pathology in schizophrenia. Sci Adv. United States; 2024;10:eadk6063.

45. Desikan RS, Ségonne F, Fischl B, et al. An automated labeling system for subdividing the human cerebral cortex on MRI scans into gyral based regions of interest. Neuroimage. 2006/03/15. 2006;31:968–980.

46. Crowley SJ, Kanel P, Roytman S, Bohnen NI, Hampstead BM. Basal forebrain integrity, cholinergic innervation and cognition in idiopathic Parkinson’s disease. Brain. England; 2024;147:1799–1808.

47. La Joie R, Perrotin A, Barré L, et al. Region-Specific Hierarchy between Atrophy, Hypometabolism, and β-Amyloid (Aβ) Load in Alzheimer&#’s Disease Dementia. J Neurosci [online serial]. 2012;32:16265 LP – 16273. Accessed at: http://www.jneurosci.org/content/32/46/16265.abstract.

48. Tremblay C, Abbasi N, Zeighami Y, et al. Sex effects on brain structure in de novo Parkinson’s disease: a multimodal neuroimaging study. Brain. England; 2020;143:3052–3066.

49. Abdelgawad A, Rahayel S, Zheng Y-Q, et al. Predicting longitudinal brain atrophy in Parkinson’s disease using a Susceptible-Infected-Removed agent-based model. Netw Neurosci (Cambridge, Mass). United States; 2023;7:906–925.

50. Alexander-Bloch AF, Shou H, Liu S, et al. On testing for spatial correspondence between maps of human brain structure and function. Neuroimage. United States; 2018;178:540–551.

51. Beck CW, Bliwise NG. Interactions are critical. CBE Life Sci. Educ. United States; 2014. p. 371–372.

52. Hall H, Reyes S, Landeck N, et al. Hippocampal Lewy pathology and cholinergic dysfunction are associated with dementia in Parkinson’s disease. Brain [online serial]. 2014;137:2493–2508. Accessed at: 10.1093/brain/awu193.

53. Tan C, Nawaz H, Lageman SK, et al. Cholinergic Nucleus 4 Degeneration and Cognitive Impairment in Isolated Rapid Eye Movement Sleep Behavior Disorder. Mov Disord. United States; 2023;38:474–479.

54. Hepp DH, Foncke EMJ, Berendse HW, et al. Damaged fiber tracts of the nucleus basalis of Meynert in Parkinson’s disease patients with visual hallucinations. Sci Rep [online serial]. 2017;7:10112. Accessed at: 10.1038/s41598-017-10146-y.

55. Delgado-Álvarez A, Matias-Guiu JA, Delgado-Alonso C, et al. Cognitive Processes Underlying Verbal Fluency in Multiple Sclerosis. Front Neurol. Switzerland; 2020;11:629183.

56. Bergamino M, Keeling EG, Mishra VR, Stokes AM, Walsh RR. Assessing White Matter Pathology in Early-Stage Parkinson Disease Using Diffusion MRI: A Systematic Review. Front Neurol [online serial]. 2020;11. Accessed at: https://www.frontiersin.org/journals/neurology/articles/10.3389/fneur.2020.00314.

57. Kamiya K, Hori M, Aoki S. NODDI in clinical research. J Neurosci Methods [online serial]. 2020;346:108908. Accessed at: https://www.sciencedirect.com/science/article/pii/S0165027020303319.

58. Brysbaert M. How Many Participants Do We Have to Include in Properly Powered Experiments? A Tutorial of Power Analysis with Reference Tables. J Cogn. England; 2019;2:16.

