## Supplementary Information for "Central cholinergic white matter pathways in prodromal and early manifest Lewy body disease"

**Reconstructing the white matter pathways of the NbM**

The DWI scans in the HCP dataset were acquired with the following main parameters: 1.05mm isotropic voxels, 132 transversal slices acquired in interleaved order without a gap, phase encoding applied along the anterior–posterior direction, two shells with b-values = 1000, 2000 s/mm^2^, repetition time/echo time = 7000/71.2 ms, 65 unique diffusion gradient directions and 6 b0 images were obtained for each phase encoding direction pair (AP and PA pairs). The total scanning time for the dMRI protocol was about 40 min.

The dMRI preprocessing was performed as described by Glasser and colleagues^1^, based on the updated diffusion pipeline (v3.19.0), including basic preprocessing, distortion correction, eddy current correction, motion correction, gradient nonlinearity correction, and registration of the mean b0 image to native T1w images with FSL’s FLIRT BBR + bbregister and transformation of diffusion images. The brain mask of each subject is based on FreeSurfer segmentation. Nonlinear transformations to standard space (MNI152) were obtained using the respective T1-weighted images using FSL’s FNIRT^2,3^. Next, estimation of the diffusion parameters was carried out using FSL’s BedpostX in a ball-and-sticks model for each voxel^4^ , considering three fibres modelled per voxel.

Probabilistic tractography was guided by several regions of interest based on previous studies^5–7^. We used masks of the cingulum and external capsule from the Johns Hopkins University white matter atlas (distributed with FSL)^8^ as waypoints for the medial and lateral tracts, respectively. Stop masks localized posterior and lateral to the NbM were also used to prevent fibres from the medial pathway running posteriorly into the fornix or merging laterally into the lateral pathway. Furthermore, based on previous human post-mortem research^9^, a retrosplenial cortical mask was used as the final stop mask for the medial pathway, while a mask of the lateral pathway’s cortical targets was used as the final stop mask for the lateral pathway^10^. In addition, we used several exclusion masks for each pathway, namely, mask of the brainstem from the Harvard-Oxford atlas in FSL^2^, mask of the anterior commissure from FSL’s XTRACT^11^, and masks of the contralateral hemispheres and corpus callosum. For each pathway we also used the other pathway’s main waypoint as an exclusion mask (for example, the cingulum was used as an exclusion mask in the reconstruction of the lateral pathway).

The resulting streamline density map of the probabilistic fibre tracking of each HCP participant was binarized, and group-level unilateral templates of the lateral and medial pathways were created by including all voxels that were included in at least 70% of the HCP participants’ binarized maps. This threshold was chosen based on visual inspection of the resulting pathways and previous works^6,7^. Lastly, we added the two unilateral templates of each pathway to create bilateral medial/lateral pathway binarized templates (**Figure 1a**).

**Extraction of microstructure indices**

Prior to running PCA, we first z-transformed each metric for each pathway and inverted the MD’s z-scores, so that both higher FA and higher MD represent higher integrity. We then ran the PCA on each pathway’s z-scored metrics and used the resulting first principal component (PC1) for each pathway (explaining 85% and 81% of the variance for the lateral and medial pathways, respectively) as the microstructural integrity measure for that pathway in all subject-level analyses.

Also, to note, the extent of outlier dMRI slices detected by EDDY among the OPDC participants was weakly associated with the different diffusion metrics of the NbM pathways (r=0.00-0.16, controlled for age and sex).

**NbM grey matter (GM) volume assessment**

The T1-weighted images were resampled, corrected for bias field inhomogeneities, affine-registered, and segmented into brain tissue classes (i.e., grey matter (GM), white matter (WM), and cerebrospinal fluid (CSF)). Further preprocessing included local tissue intensity transformation, partial volume estimation, and spatial normalization to standard MNI space using DARTEL. The spatially normalized images were then “modulated” by multiplying the voxel values with the Jacobian determinant (i.e., linear and non-linear components) derived from the spatial normalization. This allows the extraction of the absolute amount of tissue (e.g. "volume" of grey matter)^12^. GM volumes for further analyses were extracted from the final preprocessed GM volume maps, i.e., the modulated, normalized, partial volume corrected GM images of each participant.

**Disease-epicenter analysis**

NbM-cortical structural connectivity profile

To create the healthy NbM-cortical structural connectivity profile we again ran the probabilistic fibre tracking as detailed above using the ProbtracxkX2’s network mode (--network option) to quantify the number of streamlines seeded from either the left or right NbM and 34 cortical regions in each hemisphere using the Desikan-Killiany atlas in MNI space^13^. Streamlines seeded from the NbM to the cortical regions were classified to the lateral or medial NbM pathways as suggested and described previously^5,9,10^. Specifically, from the medial pathway streamlines to medial orbitofrontal, cingulate, and retrosplenial cortices were accounted for, while from the lateral pathway streamlines from dorsal and lateral frontal cortices, superior, inferior and medial parietal cortices, superior, middle and inferior temporal gyri, inferotemporal cortex, parahippocampal gyrus, occipital cortices, frontoparietal opercular cortices, and insula were accounted for (see Supplementary Figure 1 for graphical illustration). Each cortical ROI’s streamlines count for each HCP participant was weighted/normalized by dividing the number of streamlines by the number of voxels in this ROI as well as the total number of valid streamlines seeded from the NbM. The mean value of the weighted streamlines count for each of the 68 cortical regions was then calculated across the 176 HCP participants to create the healthy NbM-cortical structural connectivity profile. Regions with 0 streamlines seeded in the NbM in specific participants were not accounted for when averaging across participants.

Atrophy patterns (w-score maps)

To create w-score maps, voxelwise linear regression analysis was first performed in the OPDC healthy control group between the preprocessed GM maps created with CAT12 and age, sex, TIV, and IQR using SPM. Then, individual w-score maps were computed for each iRBD or PD patient using the following formula: w-score = [(patient’s raw value) − (value expected in the control group for the patient's covariates)]/SD of the residuals in controls. From the w-score map of each patient, we extracted the mean w-scores for the 68 cortical regions in the Desikan-Killiany atlas^13^, and then averaged the scores of each region across patients in either the iRBD or PD groups to create the syndrome-specific (i.e., iRBD or PD) cortical atrophy profile (**Figure 5a**).

Spatial correlation

Spearman’s rank correlation was used to correlate the spatial patterns of the healthy NbM connectivity profile with each patient group’s cortical atrophy pattern. Since the intrinsic spatial smoothness in two given brain maps may inflate the significance of their spatial correlation, we assessed statistical significance of these correlations using spin permutation tests^14,15^. This framework generates null models of overlap between cortical maps by projecting the spatial coordinates of cortical data onto the surface spheres, applying randomly sampled rotations (10,000 repetitions), and reassigning connectivity values. The original correlation coefficients are then compared against the null distributions determined by the sampling of correlation coefficients comparing spatially permuted cortical maps. *P*_spin_<0.05 considered statistically significant.

**Supplementary Table 1.** MoCA and verbal fluency data availability for the patient groups at baseline and follow-up visits

| **Characteristics** | **PD** | **iRBD** |
| --- | --- | --- |
| Baseline visit | MoCA: N=73  Verbal fluency: N=72 | MoCA: N=67  Verbal fluency: N=67 |
| Visit 2 | MoCA: N=69  Verbal fluency: N=69 | MoCA: N=60  Verbal fluency: N=61 |
| Visit 3 | MoCA: N=59  Verbal fluency: N=59 | MoCA: N=51  Verbal fluency: N=50 |
| Visit 4 | MoCA: N=46  Verbal fluency: N=46 | MoCA: N=42  Verbal fluency: N=42 |
| Visit 5 | MoCA: N=31  Verbal fluency: N=31 | MoCA: N=33  Verbal fluency: N=33 |
| Visit 6 | MoCA: N=18  Verbal fluency: N=18 | MoCA: N=26  Verbal fluency: N=26 |
| Visit 7 | MoCA: N=11  Verbal fluency: N=11 | MoCA: N=11  Verbal fluency: N=11 |
| Visit 8 | MoCA: N=3  Verbal fluency: N=3 | MoCA: N=6  Verbal fluency: N=6 |
| Visit 9 | MoCA: N=3  Verbal fluency: N=3 | MoCA: N=2  Verbal fluency: N=2 |

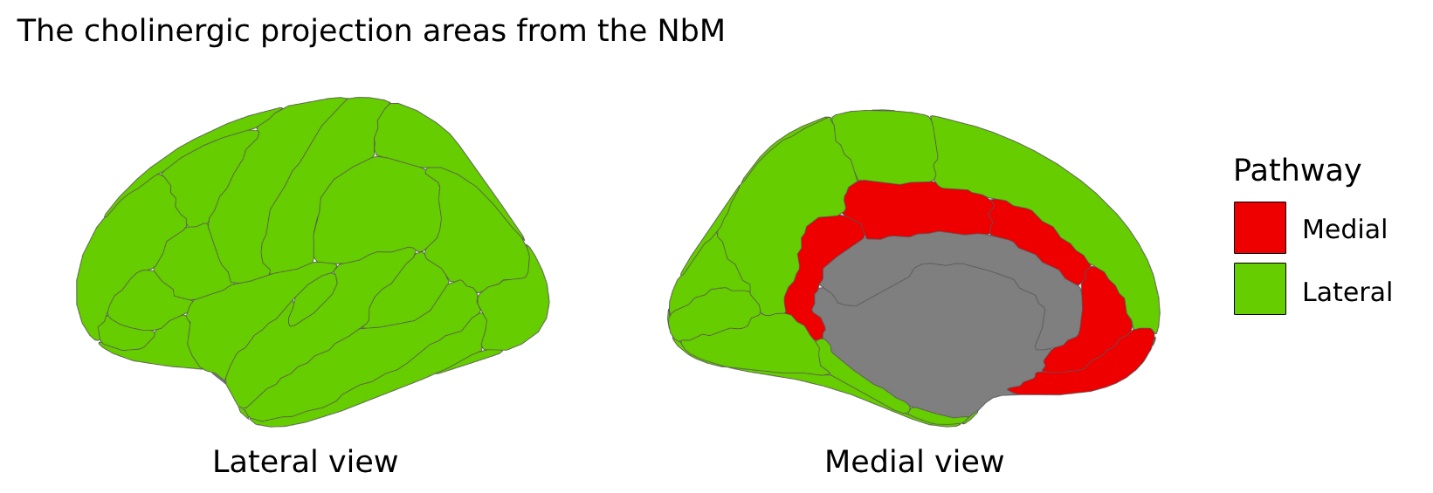

**Supplementary Figure 1.** The cholinergic projection areas from the NbM. NbM cortical projection areas used for streamlines counts based on the white matter pathways of the NbM described by Selden et al. (1998). The lateral pathway’s projection area (green) includes streamlines from dorsal and lateral frontal cortices, superior, inferior and medial parietal cortices, superior, middle and inferior temporal gyri, inferotemporal cortex, parahippocampal gyrus, occipital cortices, frontoparietal opercular cortices, and insula. The medial pathway’s projection area (red) includes streamlines from medial orbitofrontal, cingulate, and retrosplenial cortices. The open-source ggseg package in R was used for the schematic illustration.

1.	Glasser MF, Sotiropoulos SN, Wilson JA, et al. The minimal preprocessing pipelines for the Human Connectome Project. *Neuroimage*. 2013;80:105-124. doi:10.1016/j.neuroimage.2013.04.127

2.	Jenkinson M, Beckmann CF, Behrens TEJ, Woolrich MW, Smith SM. FSL. *Neuroimage*. 2012;62(2):782-790. doi:https://doi.org/10.1016/j.neuroimage.2011.09.015

3.	Andersson JLR, Jenkinson M, Andersson JLR. Non-linear optimisation FMRIB Technial Report TR 07 JA 1. In: ; 2007. https://api.semanticscholar.org/CorpusID:12921662

4.	Behrens TEJ, Berg HJ, Jbabdi S, Rushworth MFS, Woolrich MW. Probabilistic diffusion tractography with multiple fibre orientations: What can we gain? *Neuroimage*. 2007;34(1):144-155. doi:https://doi.org/10.1016/j.neuroimage.2006.09.018

5.	Mesulam M-M. Cholinergic circuitry of the human nucleus basalis and its fate in Alzheimer’s disease. *J Comp Neurol*. 2013;521(18):4124-4144. doi:10.1002/cne.23415

6.	Nemy M, Cedres N, Grothe MJ, et al. Cholinergic white matter pathways make a stronger contribution to attention and memory in normal aging than cerebrovascular health and nucleus basalis of Meynert. *Neuroimage*. 2020;211:116607. doi:10.1016/j.neuroimage.2020.116607

7.	Schumacher J, Ray NJ, Hamilton CA, et al. Cholinergic white matter pathways in dementia with Lewy bodies and Alzheimer’s disease. *Brain*. 2022;145(5):1773-1784. doi:10.1093/brain/awab372

8.	MRI Atlas of Human White Matter. *AJNR Am J Neuroradiol*. 2006;27(6):1384-1385.

9.	Selden NR, Gitelman DR, Salamon-Murayama N, Parrish TB, Mesulam MM. Trajectories of cholinergic pathways within the cerebral hemispheres of the human brain. *Brain*. 1998;121 ( Pt 1:2249-2257. doi:10.1093/brain/121.12.2249

10.	Crowley SJ, Kanel P, Roytman S, Bohnen NI, Hampstead BM. Basal forebrain integrity, cholinergic innervation and cognition in idiopathic Parkinson’s disease. *Brain*. 2024;147(5):1799-1808. doi:10.1093/brain/awad420

11.	Warrington S, Bryant KL, Khrapitchev AA, et al. XTRACT - Standardised protocols for automated tractography in the human and macaque brain. *Neuroimage*. 2020;217:116923. doi:https://doi.org/10.1016/j.neuroimage.2020.116923

12.	Good CD, Johnsrude IS, Ashburner J, Henson RNA, Friston KJ, Frackowiak RSJ. A Voxel-Based Morphometric Study of Ageing in 465 Normal Adult Human Brains. *Neuroimage*. 2001;14(1):21-36. doi:https://doi.org/10.1006/nimg.2001.0786

13.	Desikan RS, Ségonne F, Fischl B, et al. An automated labeling system for subdividing the human cerebral cortex on MRI scans into gyral based regions of interest. *Neuroimage*. 2006;31(3):968-980. doi:10.1016/j.neuroimage.2006.01.021

14.	Alexander-Bloch AF, Shou H, Liu S, et al. On testing for spatial correspondence between maps of human brain structure and function. *Neuroimage*. 2018;178:540-551. doi:10.1016/j.neuroimage.2018.05.070

15.	Larivière S, Rodríguez-Cruces R, Royer J, et al. Network-based atrophy modeling in the common epilepsies: A worldwide ENIGMA study. *Sci Adv*. 2020;6(47). doi:10.1126/sciadv.abc6457
